# Excess weight increases the risk for neurological and neuropsychiatric symptoms in post-COVID-19 condition: A systematic review and meta-analysis

**DOI:** 10.1101/2024.11.20.24317635

**Authors:** Débora Barbosa Ronca, Larissa Otaviano Mesquita, Dryelle Oliveira, Ana Cláudia Morais Godoy Figueiredo, Jun Wen, Manshu Song, Kênia Mara Baiocchi de Carvalho

**Affiliations:** Graduate Program of Public Health, Faculty of Health Sciences, University of Brasília; Brasília, Federal District, Brazil; Health Department of Federal District, Brasília, Federal District, Brazil; Graduate Program of Human Nutrition, Faculty of Health Sciences, University of Brasília, Brasília, Federal District, Brazil; Superior School of Health Sciences, Brasília, Federal District, Brazil; Faculty of Hospitality and Tourism Management, Macau University of Science and Technology, Macau SAR, China; School of Medical and Health Sciences, Edith Cowan University, Perth, Western Australia, Australia

**Keywords:** Post-COVID-19 condition, Neurological symptoms, Neuropsychiatric symptoms, Obesity, Systematic review

## Abstract

**Background/purpose**: Excess weight has been identified as a potential risk factor for the development of post-COVID-19 condition (PCC). This review investigates whether excess weight increases the risk of neurological and neuropsychiatric symptoms associated with PCC. **Methods**: Studies published up to July 2023 were searched independently across eight electronic databases to evaluate the risk of developing neurological and neuropsychiatric symptoms more than 12 weeks post-infection between exposure and controls groups (excess weight *vs.* normal weight; obesity *vs.* non-obesity). Meta-analyses were conducted under a random-effects model. **Results:** Of the 10,122 abstracts screened, 18 studies (*n* = 139,091 adults) met the inclusion criteria and reported PCC symptoms according to nutritional status. These studies included 79,050 individuals with excess weight *vs* 57,926 normal-weight individuals and 30,694 individuals with obesity *vs* 107,612 non-obese individuals. The presence of excess weight in PCC significantly increased the risk of depression (RR = 1.21; 95% CI: 1.03–1.42), headache (RR = 1.21; 95% CI: 1.09–1.35), memory issues (RR = 1.43; 95% CI: 1.24–1.65), sleep disturbance (RR = 1.31; 95% CI: 1.16–1.48), and vertigo (RR = 1.21; 95% CI: 1.04– 1.41). Obesity significantly increased the risk of headache (RR = 1.41; 95% CI: 1.34– 1.49), smell disorder (RR = 1.15; 95% CI: 1.09–1.21), taste disorder (RR = 1.21; 95% CI: 1.07–1.36), and vertigo (RR = 1.44; 95% CI: 1.35–1.53). **Conclusions:** Excess weight or obesity increases the risk of experiencing neuro-symptoms related to PCC. Individuals with these conditions urgently need enhanced personalized care management in current post-pandemic context.

## Introduction

A growing body of evidence suggests that a subset of COVID-19 survivors develop persistent, debilitating symptoms and may face a long road to complete recovery[1–4]. These symptoms have been shown to affect multiple organ systems, as evidenced by respiratory, cardiovascular, neurological, and mental health manifestations. This emergent condition has several names: long-COVID; post-acute sequelae of SARS-CoV-2 syndrome; and, as per the World Health Organization (WHO), post-COVID-19 condition (PCC). Although PCC has no uniform definition in the literature, it is generally described as involving persistent symptoms or new symptom onset, typically 12 weeks from the acute phase of COVID-19[5, 6], presenting a novel challenge to healthcare systems[7].

To date, no comprehensive effective treatment has been recognized for PCC, and the common strategies are based on symptoms relief[8]. Among PCC’s range of symptoms, long-term COVID-19 neurological and neuropsychiatric manifestations are of particular interest given the higher incidence of nervous system related sequalae’s after post-viral epidemics[9]. Prior reviews revealed that smell and taste disorders, headache, sleep disturbance, anxiety and depression were among the most commonly reported persistent symptoms in the general population. However, these studies only follow participants for a few weeks post-COVID infection and do not meet the criteria for PCC[10–13]. The severity of the COVID-19 acute phase has been associated with the development of long-term COVID-19 neuropsychiatric symptoms[14]. The body mass index (BMI) has been regarded as a potential risk factor for PCC, although the association of excess weight (EW) with the development of specific neurological and neuropsychiatric symptoms remains unclear[15, 16]. Moreover, studies that reported the prevalence of persistent neurological and neuropsychiatric symptoms up to one year after COVID-19 onset exhibited considerable heterogeneity and did not perform subgroups analyses according to nutritional status[2, 17]. An increased number of PCC symptoms that longer persist may be experienced by individuals with EW and might be associated to worsen health and poor quality of life[18].

It is noteworthy that EW is a chronic suboptimal heath condition characterized by excessive fat deposits and is considered a global public health problem. It represents a major risk factor for other chronic conditions and encompasses overweight, a suboptimal body weight that represents a risk to health, and obesity, a metabolic disease[19, 20]. A complex yet costly care of individuals with EW is required, with appropriate multidisciplinary, long-term support[19]. In 2020, the overweight and obesity pandemic collided with the COVID-19 pandemic. These conditions’ joint negative effects have increased the incidence of related diseases, leading to potentially adverse clinical and social consequences[21–23]. Obesity was found to be associated with poor COVID-19 outcomes, such as hospital admissions, intensive care admissions, and lethality rates[24–26]. Although the COVID-19 pandemic has come under control, the multisystemic nature of this disease and its long-term impacts have yet to be elucidated[10, 27].

Although there is literature on the most commonly reported neurological and neuropsychiatric symptoms of PCC, few studies reported the risk of PCC symptoms according to nutritional status and no previous review has explored the association of EW and the risk of developing specific neurological or neuropsychiatric symptoms among COVID-19 survivors. Elucidating the long-term neuro-outcomes of COVID-19, along with its suite of symptoms and high-risk populations, is urgently needed to facilitate the development of reliable and personalized care management strategies. Therefore, this systematic review addresses whether EW increases the risk of developing specific persistent neurological or neuropsychiatric symptoms among COVID-19 survivors.

## Materials and Methods

This study followed the Preferred Reporting Items for Systematic Reviews and Meta-Analyses (PRISMA) 2020 guidelines[28]. It is a subset of broad research aimed at investigating the role of EW in the development of persistent symptoms of PCC. This review was registered at the International Prospective Register of Systematic Reviews (PROSPERO) (ID: CRD42023433234).

### Search strategy and Eligibility Criteria

Two authors (DBR and LOM) searched seven electronic databases (MEDLINE, EMBASE, SCOPUS, Web of Science, VHL, Google Scholar, ProQuest) and a preprint server (medRxiv) on 3 July 2023. The search strategy was reviewed by two reviewers according to the criteria of the Peer Review of Electronic Search Strategies (PRESS) checklist[29]. The search strategy included the following terms: (“long COVID-19” OR “post-acute covid 19 syndrome” OR “COVID-19” OR “SARS-CoV-2” OR “post-COVID-19 condition” OR “long hauler”) AND (“excess of weight” OR “overweight” OR “obesity” OR “body mass index”) AND (“signs and symptoms” OR “COVID-19 sequelae” OR “headache” OR “loss of smell” OR “cognitive dysfunction” OR “anxiety” OR “depression” OR “sleep disorder” OR “brain fog).” The full search strategy is detailed in supporting information (S1 Table). Additionally, some articles were hand-searched to identify potentially eligible studies which might not be electronically retrieved, including those published in 2024.

Studies were considered eligible for inclusion if they (a) had observational designs; (b) primarily focused on adults; (c) classified focal populations based on the presence of excess weight/obesity *versus* normal weight/non-obesity, using either measured BMI or self-reported data; and (d) had a mean (or median) follow-up of at least 12 weeks after the acute phase of COVID-19. Survivors could be (a) hospitalized or non-hospitalized, (b) inpatient or outpatient, and (c) mixed population (hospitalized and outpatient) recruited in the community, outpatient clinic or heath care system.

We exclude editorials, clinical trials, reviews, opinions, books or book chapters, conference abstracts, case reports, and correspondence articles. Studies were also excluded if they included mostly pregnant or nursing women; evaluated the health effects of COVID-19; assessed excess weight as a risk factor for acute-phase severity or mortality; or evaluated long-term sequelae of COVID-19 in a specific population (e.g., individuals with specific comorbidities). Additionally, studies were excluded if they investigated clusters of symptoms rather than specific symptoms or if they lacked a control group (i.e., COVID-19 survivors who did not report persistent symptoms).

### Screening Process, Study Selection, and Data Extraction

The same two authors (DBR and LOM) independently screened the titles and abstracts of studies obtained from database searches and removed duplicates. The Rayyan app[30], a semi-automation tool, was used to streamline the review process. This was followed by a full-text review of the retained articles to ensure they met eligibility criteria. Fig 1 presents the flowchart for the included studies.

**Fig 1:**
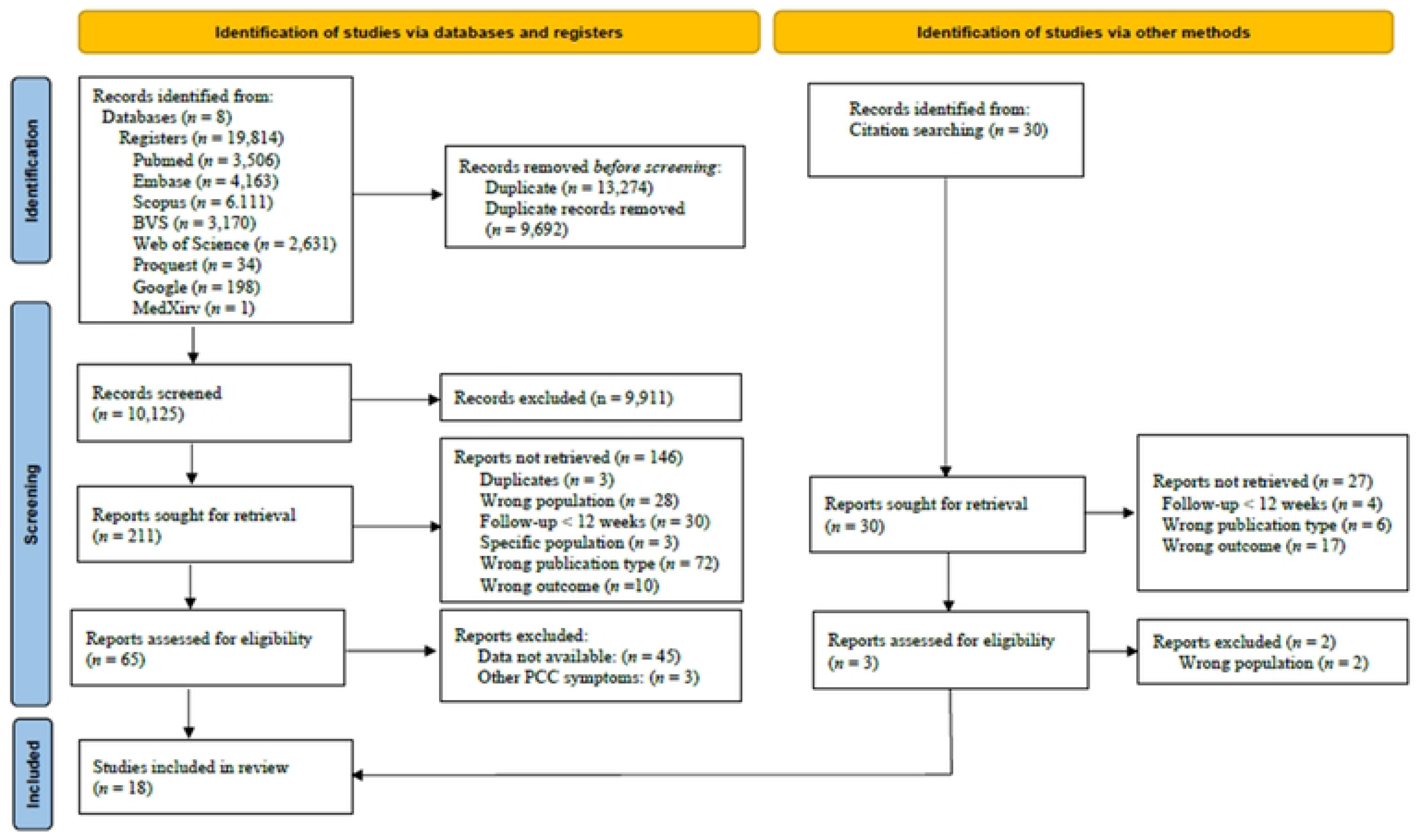
Flow diagram of included studies.

The study population of interest comprised COVID-19 survivors. Exposure was defined as EW or obesity based on BMI measurement, data from electronic medical records, or self-reported information. Two exposure groups were defined according to WHO guidelines[31]: 1) an excess weight group (BMI ≥25 kg/m^2^) and 2) an obesity group (BMI ≥30 kg/m^2^). Both Asian and Caucasian populations were included in the analysis, with corresponding BMI cut-offs applied: for Asian-Pacific populations, the thresholds were BMI ≥23 kg/m^2^ for EW and BMI ≥25 kg/m^2^ for obesity, as previously defined in the included studies[32, 33].

We adopted WHO’s definition of PCC as outlined by Soriano *et al*. (2021)[5]. Symptoms were measured with predefined questionnaires/scales or reported during interviews (in person, by phone, or online). Due to varying definitions illness onset (baseline), we accepted definitions that included 12 weeks from the onset of COVID-19 symptoms, COVID-19 diagnosis, hospital admission or discharge after the acute phase. For the follow-up period, we considered studies where the sample median (or mean) of persistent symptoms was reported at least 12 weeks (84 days) after the baseline, with a minimum interquartile range (IQR) of 10 weeks (or a standard deviation (SD) of ±24 days) from baseline. This cut-off was necessary to include studies evaluating long-term sequelae of COVID-19. Given the varied terminology used for symptoms across studies, we re-grouped symptoms into neurological and neuropsychiatric categories as shown in Fig 2.

**Fig 2:**
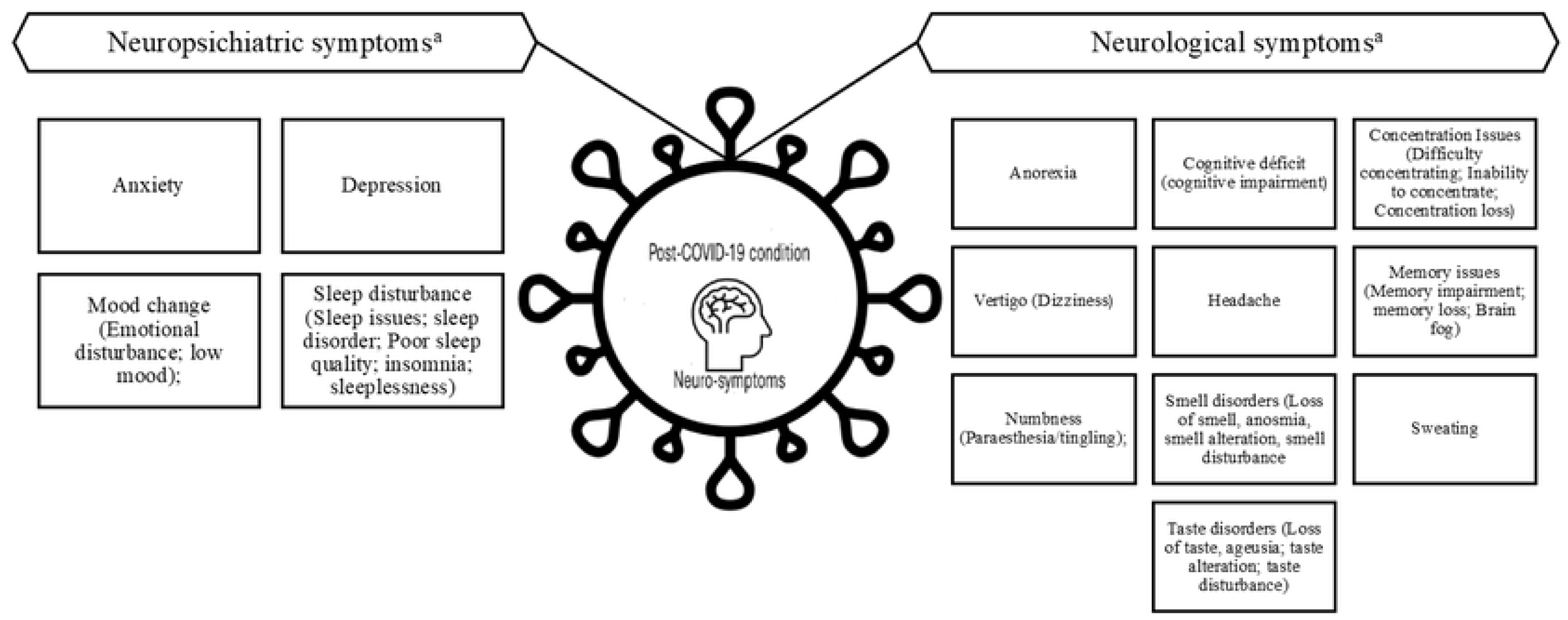
Reported neurological and neuropsychiatric symptoms related to Post-COVID-19 condition (PCC) ^a^ Neurological and neuropsychiatric reported symptoms were grouped into one category with synonyms presented in brackets.

Specific data were exported from each study into a predefined data collection form, including authors; publication year; country; aim of the study; study design; mean follow-up period; study population; exposure groups; assessment of outcome; evaluated symptoms; frequency of symptoms (*n*, %) among exposure and control groups; and the effect measure of the association of EW and the neurological/neuropsychiatric symptom. Corresponding authors of relevant articles published were contacted if any data were absent.

### Quality Assessment

The ROBINS-E tool for observational research was used to determine the risk of bias in the included studies[34]. ROBINS-E is a domain-based tool that evaluates seven domains (confounders, exposure measurement, participant selection, post-exposure interventions, missing data, outcome measurement, and selection of reported results) and then a general assessment of the study. Traffic plots with a final analysis of studies’ risk of bias, by domain, is generated. For this research, two authors (DBR and DO) initially appraised the risk of bias. Disagreements were discussed and resolved in collaboration with a third researcher (LOM). The ROBINS-E tool was not applied as a study eligibility criterion.

We assessed the certainty of evidence via the Grading of Recommendations Assessment, Development and Evaluation (GRADE) framework [35–37] regarding the risk of developing PCC-related neurological and neuropsychiatric symptoms among individuals with EW. The GRADE system measures attributes that impact the overall quality of evidence. We assessed the certainty of evidence of the outcomes that were statistically significantly associated with exposures groups and scored it accordingly (i.e., high = ≥4 points; moderate = 3 points; low = 2 points; very low = 1 point).

### Statistical Analysis

The frequency of reported symptoms and the associated risk of development were determined for EW *vs* normal weight groups and for obesity *vs* non-obesity groups, classified by BMI when available (normal weight: BMI of 18–24.9 kg/m^2^; excess weight: BMI >25 kg/m^2^; obesity: BMI >30 kg/m^2^; non-obesity group: BMI <30 kg/m^2^), taking into account different cut-off values previously defined for the Chinese population in studies[32, 33]. Each symptom was considered an individual outcome. For this purpose, we collected data of the frequency (*n*, %) of each neurological and neuropsychiatric symptoms related to PCC reported and the risk of developing persistent symptoms (as measured by the odds ratio [OR] or the adjusted OR) at follow-up in included studies.

We performed meta-analytic calculations using STATA software (SE/17). Pooled risk ratios (RRs) with their 95% confidence intervals (CIs) were computed from the raw data of the included studies. A random effects model meta-analysis was conducted to account for the statistical and methodological heterogeneity of the data. We used a two-sample binary-outcome summary dataset format with DerSimonian–Laird estimation when fewer than five studies were available[38]; when more than five studies were available, we adopted restricted maximum-likelihood estimation and to conduct pooled RR analysis. Statistical heterogeneity was assessed using the *I*^2^ statistic[39]. Forest plots were generated for all outcomes. Publication bias was examined by visual inspection of funnel plots. A value of *p* < 0.05 was considered statistically significant.

## Results

Of the 10,122 abstracts screened, 211 full-text registers were reviewed, and 65 studies were considered eligible. Data on neurological and neuropsychiatric symptoms according to individuals’ nutritional status were available for 18 studies (including one preprint[40]), all of which were entered into our meta-analysis.

Selected studies originated from 23 countries across Asia (China[32, 33], India[41], Indonesia[42], Japan[43], Malaysia[40] and Saudi Arabia[44]), Europe (Denmark[45, 46], England[47], Germany[48], Italy[49], Poland[50], Spain[51] and Switzerland[52]), and North and South America (Argentina[53], Brazil[53], Chile[53], Dominican Republic[53], Ecuador[53], Mexico[53], Panama[53], Paraguay[53], Peru[53] and United States of America[54, 55]), comprising both developed and developing countries. Among the included studies, two employed case–control design[51, 54], six involved retrospective or prospective cohorts[33, 42, 49, 50, 52, 55], and 10 involved cross-sectional studies[32, 40, 41, 44–47, 47, 48, 53].

The populations investigated varied: three studies evaluated individuals in outpatient settings[46, 52, 54], five involved hospitalized patients[32, 33, 42, 49, 51], and ten included individuals either hospitalized or outpatient settings during COVID-19 acute-phase[40, 41, 43, 44, 47, 48, 50, 53, 55]. The classification of individuals according to nutritional status also varied: four studies compared symptoms between individuals with EW and those with normal weight[32, 40, 42, 44], five studies evaluated obesity *versus* non-obesity[33, 48–51], eight studies presented data for individuals with normal weight, overweight and obesity[41, 45–47, 52–55], and one study only provided the mean BMI of the sample without specifying the number of individuals in exposure and control groups[43]. In total, our meta-analysis included 79,050 people with EW and 30,694 with obesity in exposure groups, while 57,926 normal weight individuals and 107,612 non-obese individuals comprised control groups. Sample sizes ranged from 32[54] to 78,566[47] COVID-19 survivors. Two Chinese studies applied different cut-off points for nutritional status classification (EW as BMI ≥23 kg/m^2^; obesity as BMI ≥25 kg/m^2^)[32, 33]. Exposure was assessed via anthropometric measures[32, 33, 41, 50, 53, 54], electronic medical record data[42, 46, 49, 51], and self-reported comorbidity/BMI[40, 43–45, 47, 52, 55]. PCC neurological and neuropsychiatric symptoms were evaluated using questionnaires[33, 41, 43–47, 50–52, 54], validated scales[32, 40, 48, 49, 51, 54], or self-report instruments[42, 47, 48, 53, 55]. The mean time from COVID-19 onset (baseline) to long-term symptom assessment was 25.8 weeks (range: 12 weeks[42, 44, 46, 47, 49] to 52 weeks [one year][32]). Table 1 outlines the features of the included articles.

**Table 1.**
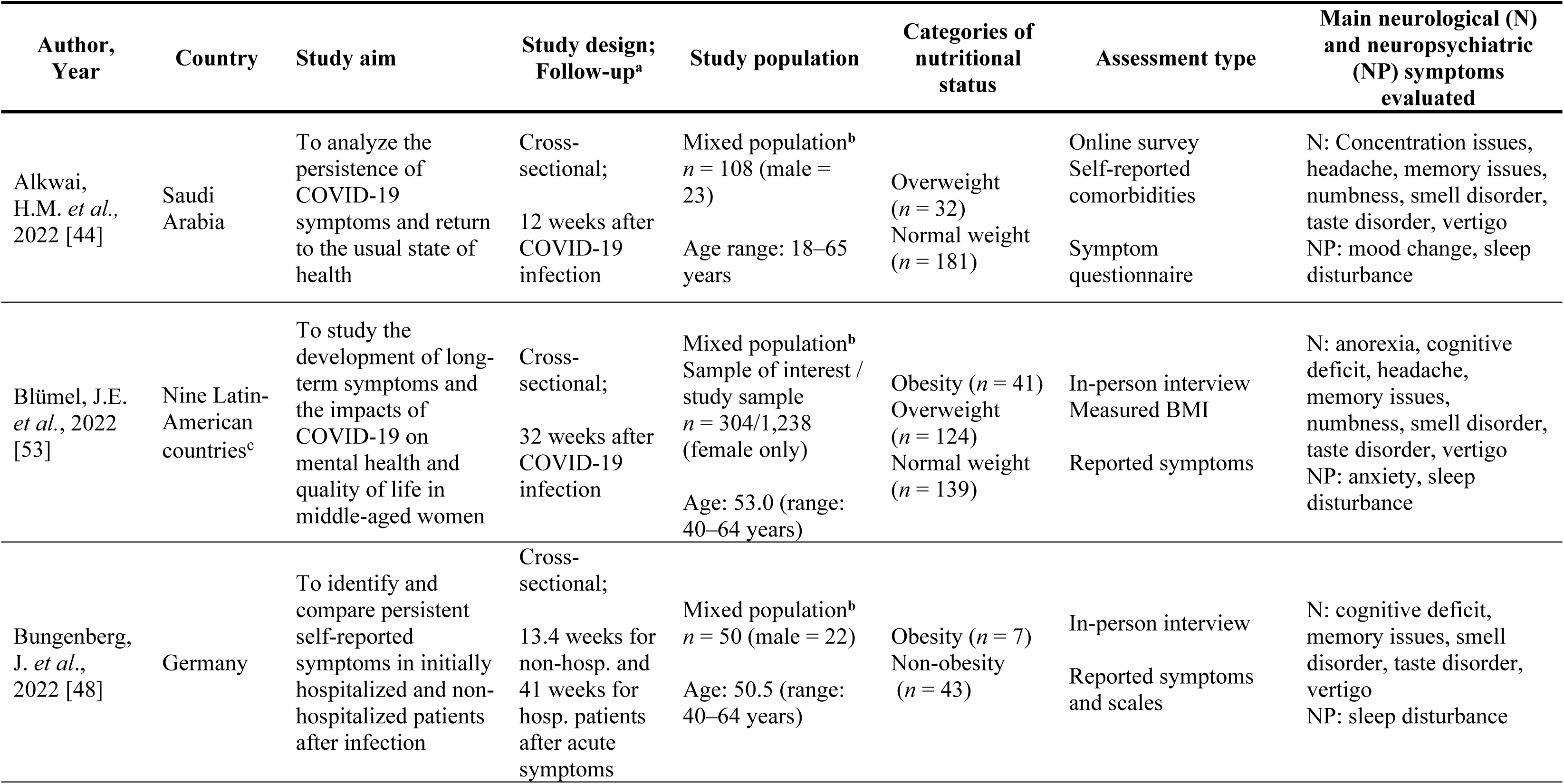
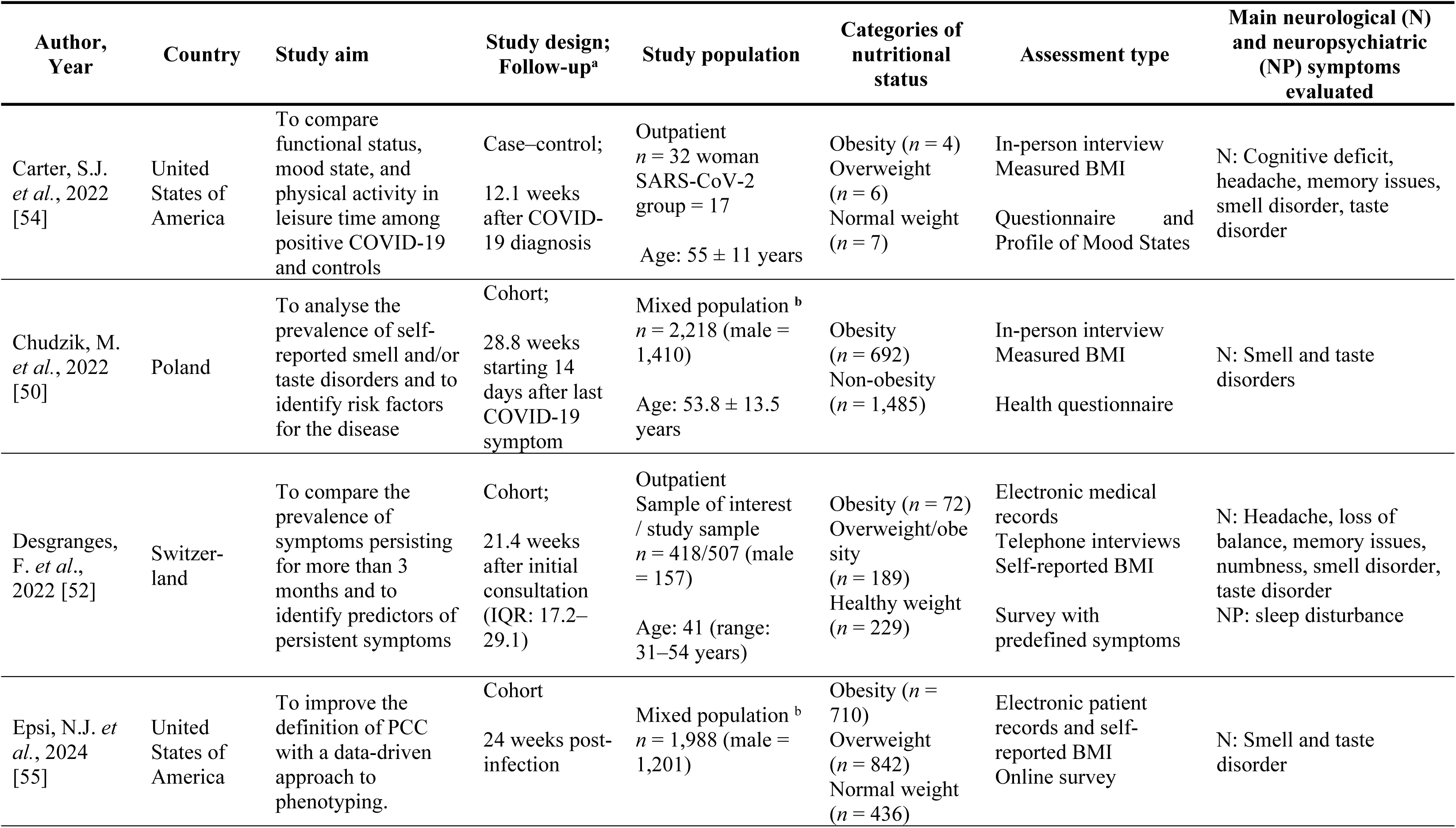
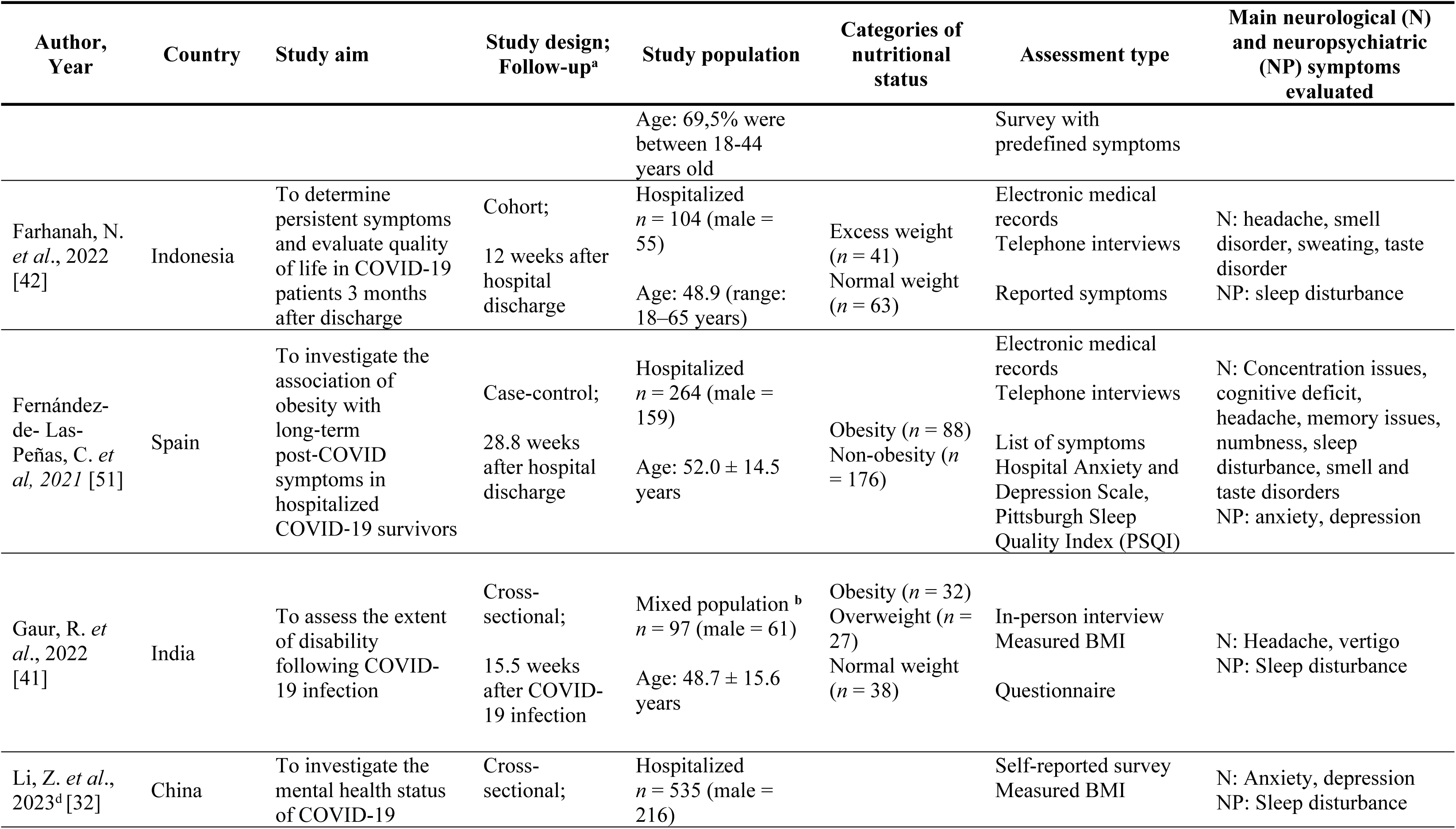
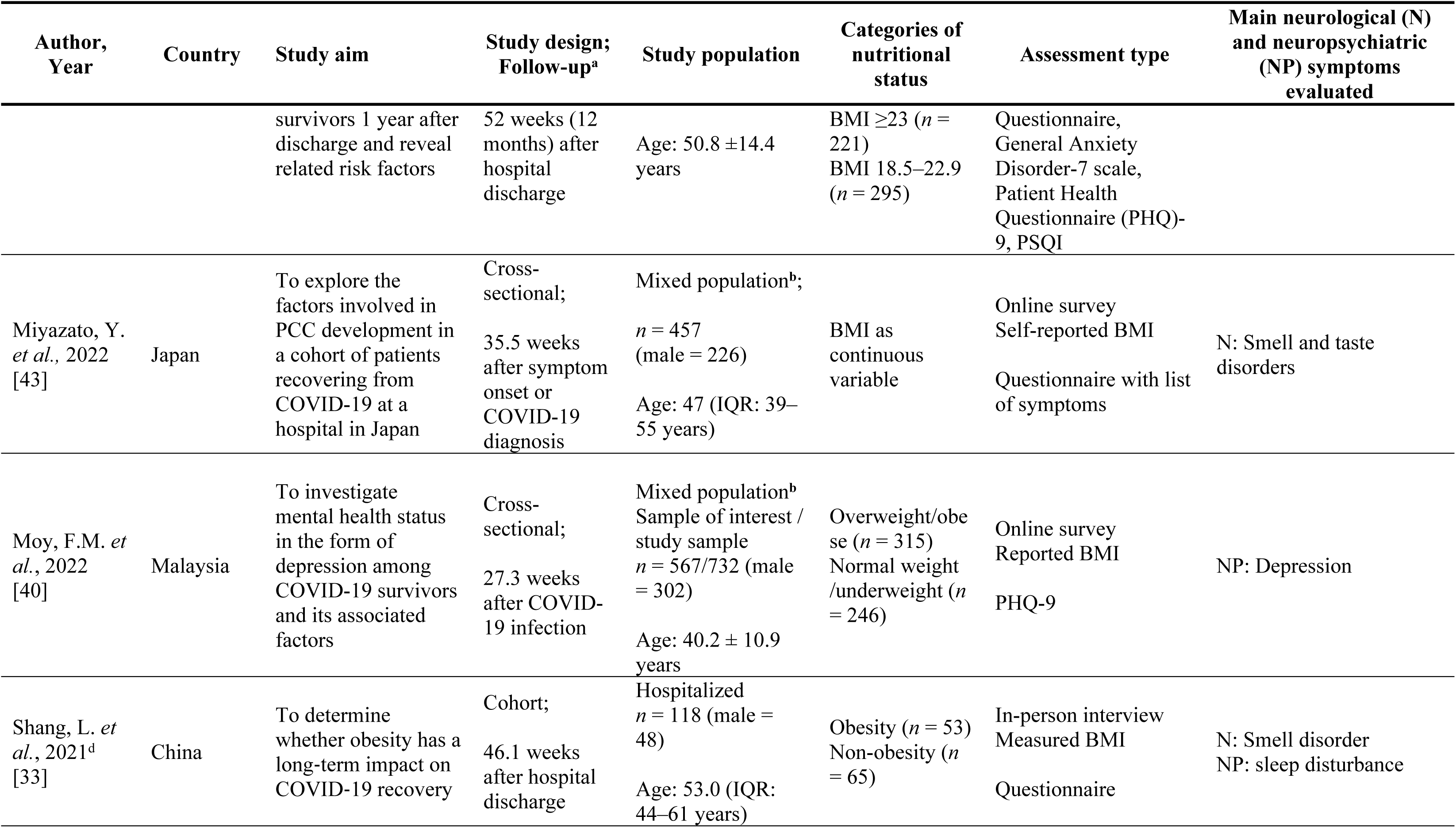
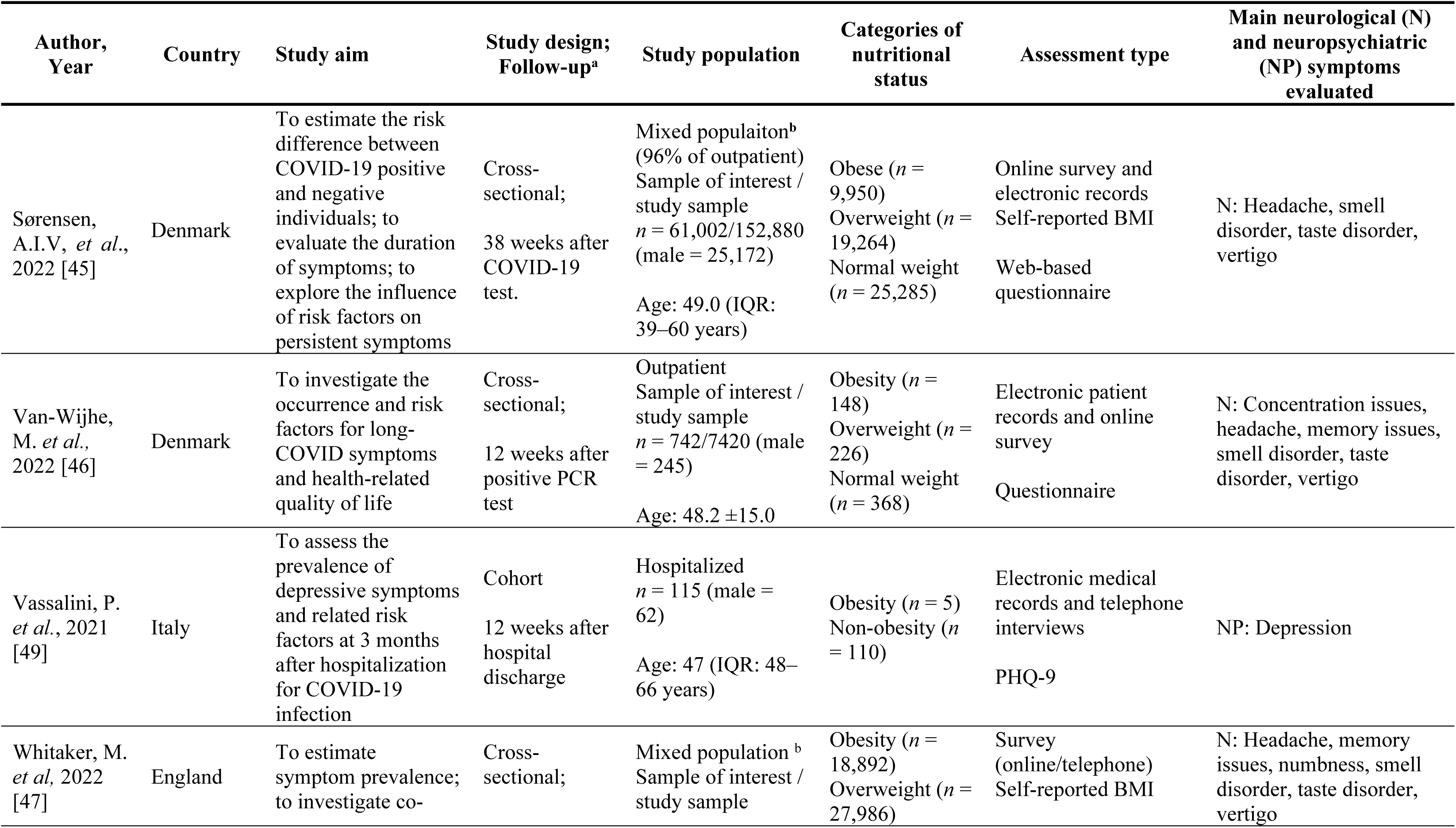
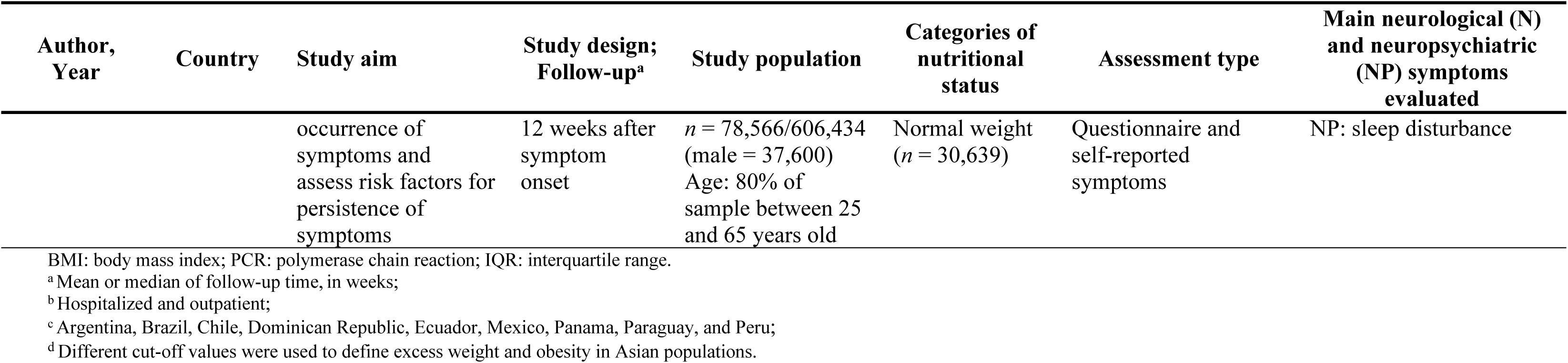
Characteristics of included studies.

| Author,<br>Year | Country | Study aim | Study design;<br>Follow-up<sup>a</sup> | Study population | Categories of<br>nutritional<br>status | Assessment type | Main neurological (N)<br>and neuropsychiatric<br>(NP) symptoms<br>evaluated |
| --- | --- | --- | --- | --- | --- | --- | --- |
| Alkwai,<br>H.M. *et al.*,<br>2022 [44] | Saudi<br>Arabia | To analyze the<br>persistence of<br>COVID-19<br>symptoms and return<br>to the usual state of<br>health | Cross-sectional;<br>12 weeks after<br>COVID-19<br>infection | Mixed population<sup>b</sup><br>*n* = 108 (male =<br>23)<br>Age range: 18–65<br>years | Overweight<br>(*n* = 32)<br>Normal weight<br>(*n* = 181) | Online survey<br>Self-reported<br>comorbidities<br>Symptom<br>questionnaire | N: Concentration issues,<br>headache, memory issues,<br>numbness, smell disorder,<br>taste disorder, vertigo<br>NP: mood change, sleep<br>disturbance |
| Blümel, J.E.<br>*et al.*, 2022<br>[53] | Nine Latin-American<br>countries<sup>c</sup> | To study the<br>development of long-term symptoms and<br>the impacts of<br>COVID-19 on<br>mental health and<br>quality of life in<br>middle-aged women | Cross-sectional;<br>32 weeks after<br>COVID-19<br>infection | Mixed population<sup>b</sup><br>Sample of interest /<br>study sample<br>*n* = 304/1,238<br>(female only)<br>Age: 53.0 (range:<br>40–64 years) | Obesity (*n* = 41)<br>Overweight<br>(*n* = 124)<br>Normal weight<br>(*n* = 139) | In-person interview<br>Measured BMI<br>Reported symptoms | N: anorexia, cognitive<br>deficit, headache,<br>memory issues,<br>numbness, smell disorder,<br>taste disorder, vertigo<br>NP: anxiety, sleep<br>disturbance |
| Bungenberg,<br>J. *et al.*,<br>2022 [48] | Germany | To identify and<br>compare persistent<br>self-reported<br>symptoms in initially<br>hospitalized and non-hospitalized patients<br>after infection | Cross-sectional;<br>13.4 weeks for<br>non-hosp. and<br>41 weeks for<br>hosp. patients<br>after acute<br>symptoms | Mixed population<sup>b</sup><br>*n* = 50 (male = 22)<br>Age: 50.5 (range:<br>40–64 years) | Obesity (*n* = 7)<br>Non-obesity<br>(*n* = 43) | In-person interview<br>Reported symptoms<br>and scales | N: cognitive deficit,<br>memory issues, smell<br>disorder, taste disorder,<br>vertigo<br>NP: sleep disturbance |
| Carter, S.J.<br>*et al.*, 2022<br>[54] | United<br>States of<br>America | To compare<br>functional status,<br>mood state, and<br>physical activity in<br>leisure time among<br>positive COVID-19<br>and controls | Case-control;<br>12.1 weeks<br>after COVID-19<br>diagnosis | Outpatient<br>*n* = 32 woman<br>SARS-CoV-2<br>group = 17<br>Age: 55 ± 11 years | Obesity (*n* = 4)<br>Overweight<br>(*n* = 6)<br>Normal weight<br>(*n* = 7) | In-person interview<br>Measured BMI<br>Questionnaire and<br>Profile of Mood States | N: Cognitive deficit,<br>headache, memory issues,<br>smell disorder, taste<br>disorder |
| Chudzik, M.<br>*et al.*, 2022<br>[50] | Poland | To analyse the<br>prevalence of self-reported smell and/or<br>taste disorders and to<br>identify risk factors<br>for the disease | Cohort;<br>28.8 weeks<br>starting 14<br>days after last<br>COVID-19<br>symptom | Mixed population <sup>b</sup><br>*n* = 2,218 (male =<br>1,410)<br>Age: 53.8 ± 13.5<br>years | Obesity<br>(*n* = 692)<br>Non-obesity<br>(*n* = 1,485) | In-person interview<br>Measured BMI<br>Health questionnaire | N: Smell and taste<br>disorders |
| Desgranges,<br>F. *et al.*,<br>2022 [52] | Switzer-<br>land | To compare the<br>prevalence of<br>symptoms persisting<br>for more than 3<br>months and to<br>identify predictors of<br>persistent symptoms | Cohort;<br>21.4 weeks<br>after initial<br>consultation<br>(IQR: 17.2–<br>29.1) | Outpatient<br>Sample of interest<br>/ study sample<br>*n* = 418/507 (male<br>= 157)<br>Age: 41 (range:<br>31–54 years) | Obesity (*n* = 72)<br>Overweight/obe<br>sity<br>(*n* = 189)<br>Healthy weight<br>(*n* = 229) | Electronic medical<br>records<br>Telephone interviews<br>Self-reported BMI<br>Survey with<br>predefined symptoms | N: Headache, loss of<br>balance, memory issues,<br>numbness, smell disorder,<br>taste disorder<br>NP: sleep disturbance |
| Epsi, N.J. *et al.*, 2024<br>[55] | United<br>States of<br>America | To improve the<br>definition of PCC<br>with a data-driven<br>approach to<br>phenotyping. | Cohort<br>24 weeks post-infection | Mixed population <sup>b</sup><br>*n* = 1,988 (male =<br>1,201) | Obesity (*n* =<br>710)<br>Overweight<br>(*n* = 842)<br>Normal weight<br>(*n* = 436) | Electronic patient<br>records and self-reported BMI<br>Online survey | N: Smell and taste<br>disorder |

| Author, Year | Country | Study aim | Study design; Follow-up<sup>a</sup> | Study population | Categories of nutritional status | Assessment type | Main neurological (N) and neuropsychiatric (NP) symptoms evaluated |
| --- | --- | --- | --- | --- | --- | --- | --- |
|  |  |  |  | Age: 69,5% were between 18-44 years old |  | Survey with predefined symptoms |  |
| Farhanah, N. *et al.*, 2022 [42] | Indonesia | To determine persistent symptoms and evaluate quality of life in COVID-19 patients 3 months after discharge | Cohort;<br>12 weeks after hospital discharge | Hospitalized<br>*n* = 104 (male = 55)<br>Age: 48.9 (range: 18–65 years) | Excess weight (*n* = 41)<br>Normal weight (*n* = 63) | Electronic medical records<br>Telephone interviews<br>Reported symptoms | N: headache, smell disorder, sweating, taste disorder<br>NP: sleep disturbance |
| Fernández-de- Las-Peñas, C. *et al.*, 2021 [51] | Spain | To investigate the association of obesity with long-term post-COVID symptoms in hospitalized COVID-19 survivors | Case-control;<br>28.8 weeks after hospital discharge | Hospitalized<br>*n* = 264 (male = 159)<br>Age: 52.0 ± 14.5 years | Obesity (*n* = 88)<br>Non-obesity (*n* = 176) | Electronic medical records<br>Telephone interviews<br>List of symptoms<br>Hospital Anxiety and Depression Scale, Pittsburgh Sleep Quality Index (PSQI) | N: Concentration issues, cognitive deficit, headache, memory issues, numbness, sleep disturbance, smell and taste disorders<br>NP: anxiety, depression |
| Gaur, R. *et al.*, 2022 [41] | India | To assess the extent of disability following COVID-19 infection | Cross-sectional;<br>15.5 weeks after COVID-19 infection | Mixed population <sup>b</sup><br>*n* = 97 (male = 61)<br>Age: 48.7 ± 15.6 years | Obesity (*n* = 32)<br>Overweight (*n* = 27)<br>Normal weight (*n* = 38) | In-person interview<br>Measured BMI<br>Questionnaire | N: Headache, vertigo<br>NP: Sleep disturbance |
| Li, Z. *et al.*, 2023<sup>d</sup> [32] | China | To investigate the mental health status of COVID-19 | Cross-sectional; | Hospitalized<br>*n* = 535 (male = 216) |  | Self-reported survey<br>Measured BMI | N: Anxiety, depression<br>NP: Sleep disturbance |
|  |  | survivors 1 year after discharge and reveal related risk factors | 52 weeks (12 months) after hospital discharge | Age: 50.8 ± 14.4 years | BMI ≥ 23 (*n* = 221)<br>BMI 18.5–22.9 (*n* = 295) | Questionnaire, General Anxiety Disorder-7 scale, Patient Health Questionnaire (PHQ)-9, PSQI |  |
| Miyazato, Y. *et al.*, 2022 [43] | Japan | To explore the factors involved in PCC development in a cohort of patients recovering from COVID-19 at a hospital in Japan | Cross-sectional;<br>35.5 weeks after symptom onset or COVID-19 diagnosis | Mixed population<sup>b</sup>;<br>*n* = 457 (male = 226)<br>Age: 47 (IQR: 39–55 years) | BMI as continuous variable | Online survey<br>Self-reported BMI<br>Questionnaire with list of symptoms | N: Smell and taste disorders |
| Moy, F.M. *et al.*, 2022 [40] | Malaysia | To investigate mental health status in the form of depression among COVID-19 survivors and its associated factors | Cross-sectional;<br>27.3 weeks after COVID-19 infection | Mixed population<sup>b</sup><br>Sample of interest / study sample<br>*n* = 567/732 (male = 302)<br>Age: 40.2 ± 10.9 years | Overweight/obese (*n* = 315)<br>Normal weight /underweight (*n* = 246) | Online survey<br>Reported BMI<br>PHQ-9 | NP: Depression |
| Shang, L. *et al.*, 2021<sup>d</sup> [33] | China | To determine whether obesity has a long-term impact on COVID-19 recovery | Cohort;<br>46.1 weeks after hospital discharge | Hospitalized<br>*n* = 118 (male = 48)<br>Age: 53.0 (IQR: 44–61 years) | Obesity (*n* = 53)<br>Non-obesity (*n* = 65) | In-person interview<br>Measured BMI<br>Questionnaire | N: Smell disorder<br>NP: sleep disturbance |
| Sørensen, A.I.V., *et al.*, 2022 [45] | Denmark | To estimate the risk difference between COVID-19 positive and negative individuals; to evaluate the duration of symptoms; to explore the influence of risk factors on persistent symptoms | Cross-sectional;<br>38 weeks after COVID-19 test. | Mixed populaiton<sup>b</sup> (96% of outpatient)<br>Sample of interest / study sample<br>$n = 61,002/152,880$ (male = 25,172)<br>Age: 49.0 (IQR: 39–60 years) | Obese ($n = 9,950$)<br>Overweight ($n = 19,264$)<br>Normal weight ($n = 25,285$) | Online survey and electronic records<br>Self-reported BMI<br>Web-based questionnaire | N: Headache, smell disorder, taste disorder, vertigo |
| Van-Wijhe, M. *et al.*, 2022 [46] | Denmark | To investigate the occurrence and risk factors for long-COVID symptoms and health-related quality of life | Cross-sectional;<br>12 weeks after positive PCR test | Outpatient<br>Sample of interest / study sample<br>$n = 742/7420$ (male = 245)<br>Age: $48.2 \pm 15.0$ | Obesity ($n = 148$)<br>Overweight ($n = 226$)<br>Normal weight ($n = 368$) | Electronic patient records and online survey<br>Questionnaire | N: Concentration issues, headache, memory issues, smell disorder, taste disorder, vertigo |
| Vassalini, P. *et al.*, 2021 [49] | Italy | To assess the prevalence of depressive symptoms and related risk factors at 3 months after hospitalization for COVID-19 infection | Cohort<br>12 weeks after hospital discharge | Hospitalized<br>$n = 115$ (male = 62)<br>Age: 47 (IQR: 48–66 years) | Obesity ($n = 5$)<br>Non-obesity ($n = 110$) | Electronic medical records and telephone interviews<br>PHQ-9 | NP: Depression |
| Whitaker, M. *et al.*, 2022 [47] | England | To estimate symptom prevalence; to investigate co- | Cross-sectional; | Mixed population <sup>b</sup><br>Sample of interest / study sample | Obesity ($n = 18,892$)<br>Overweight ($n = 27,986$) | Survey (online/telephone)<br>Self-reported BMI | N: Headache, memory issues, numbness, smell disorder, taste disorder, vertigo |
|  |  | occurrence of<br>symptoms and<br>assess risk factors for<br>persistence of<br>symptoms | 12 weeks after<br>symptom<br>onset | *n* = 78,566/606,434<br>(male = 37,600)<br>Age: 80% of<br>sample between 25<br>and 65 years old | Normal weight<br>(*n* = 30,639) | Questionnaire and<br>self-reported<br>symptoms | NP: sleep disturbance |

BMI: body mass index; PCR: polymerase chain reaction; IQR: interquartile range. <sup>a</sup> Mean or median of follow-up time, in weeks; <sup>b</sup> Hospitalized and outpatient; <sup>c</sup> Argentina, Brazil, Chile, Dominican Republic, Ecuador, Mexico, Panama, Paraguay, and Peru; <sup>d</sup> Different cut-off values were used to define excess weight and obesity in Asian populations.

**Table 2.**
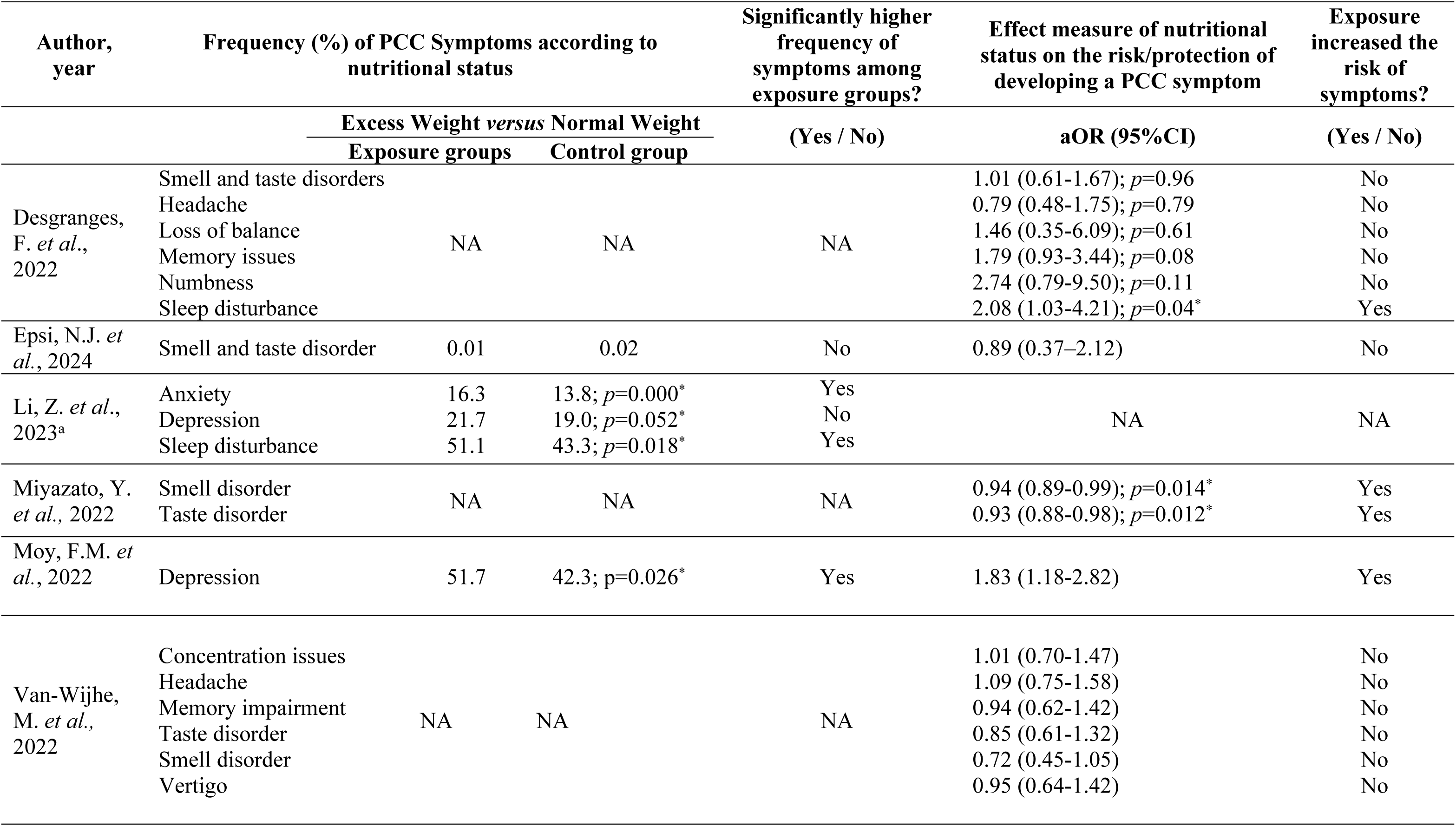
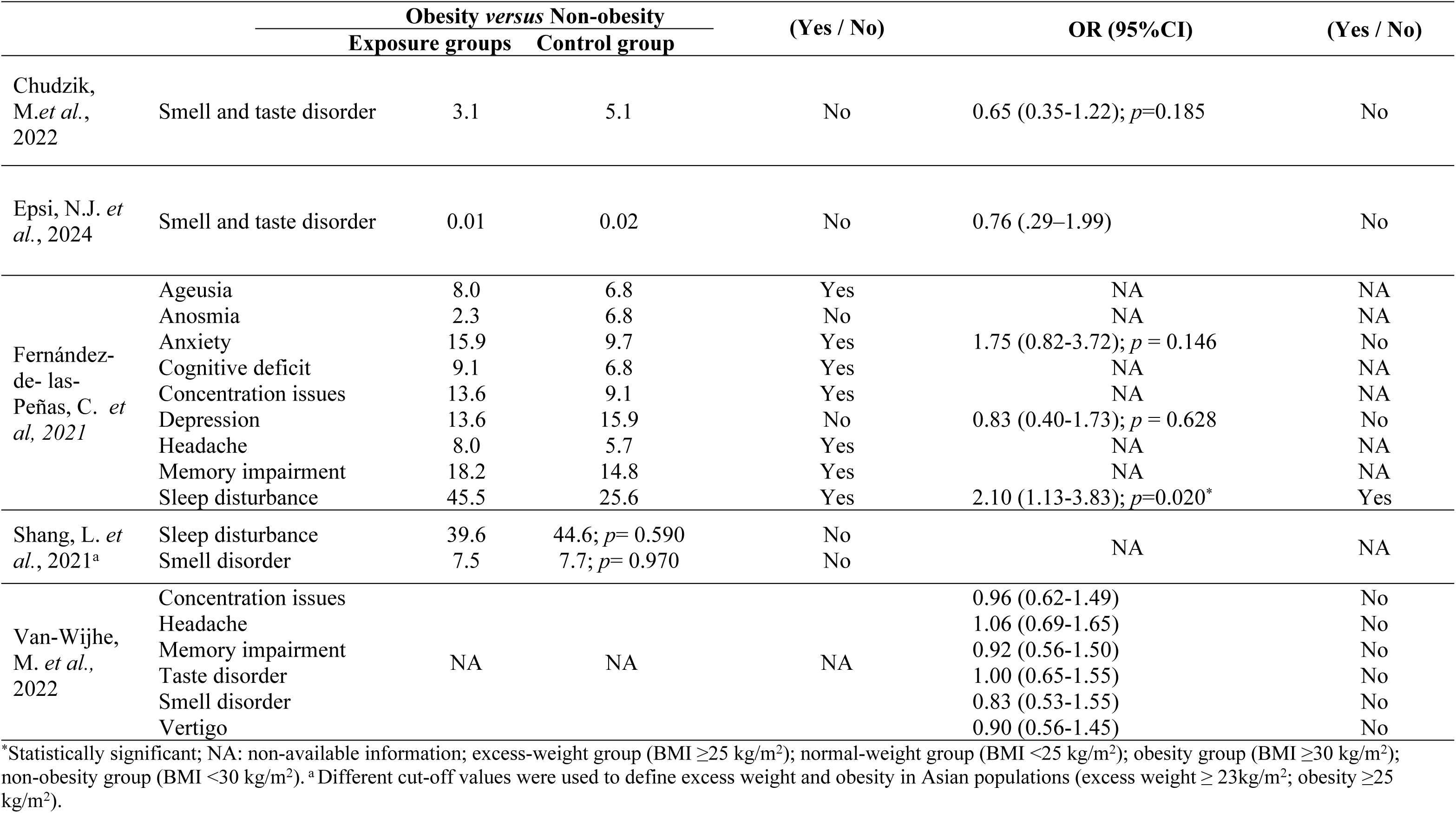
Frequency of neuropsychiatric symptoms related to post-COVID condition and the risk of their development in included studies.

### Excess Weight and the Risk of Neuropsychiatric Symptoms

We assessed the risk ratios (RRs) for the EW group (Figs 3a and 3b). Excess weight significantly increased the risk of depression (RR = 1.21; 95% CI: 1.03–1.42; *I*^2^ = 0.00), headache (RR = 1.21; 95% CI: 1.09–1.35; *I*^2^ = 41.3%), memory issues (RR = 1.43; 95% CI: 1.24–1.65; *I*^2^ = 0.00), numbness (RR = 1.37; 95% CI: 1.24–1.51; *I*^2^ = 0.00), sleep disturbance (RR = 1.31; 95% CI: 1.16–1.48; *I*^2^ = 17.83), taste disorder (RR = 1.11; 95% CI: 1.00–1.23; *I*^2^ = 53.46), and vertigo (RR = 1.21; 95% CI: 1.04–1.41; *I*^2^ = 0.00). No significant association was found between EW and other evaluated symptoms (S1 Figs 1a and 1b).

**Fig 3a.**
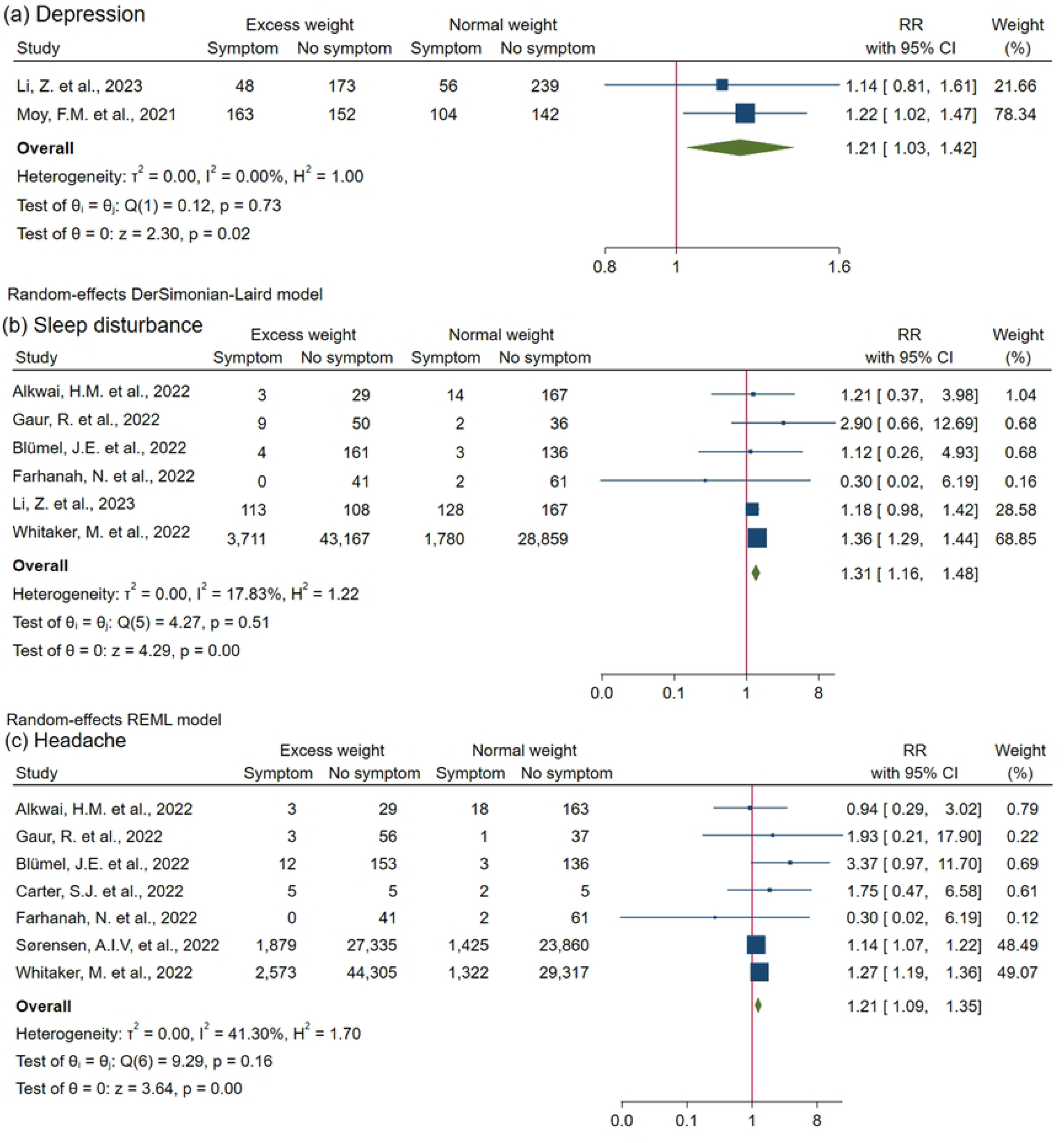
Forest plots of excess weight and risk ratio (RR) for neurological and neuropsychiatric symptoms. ^a^ Applied a different cut-off of BMI. Excess weight > 23kg/m^2.^

**Fig 3b.**
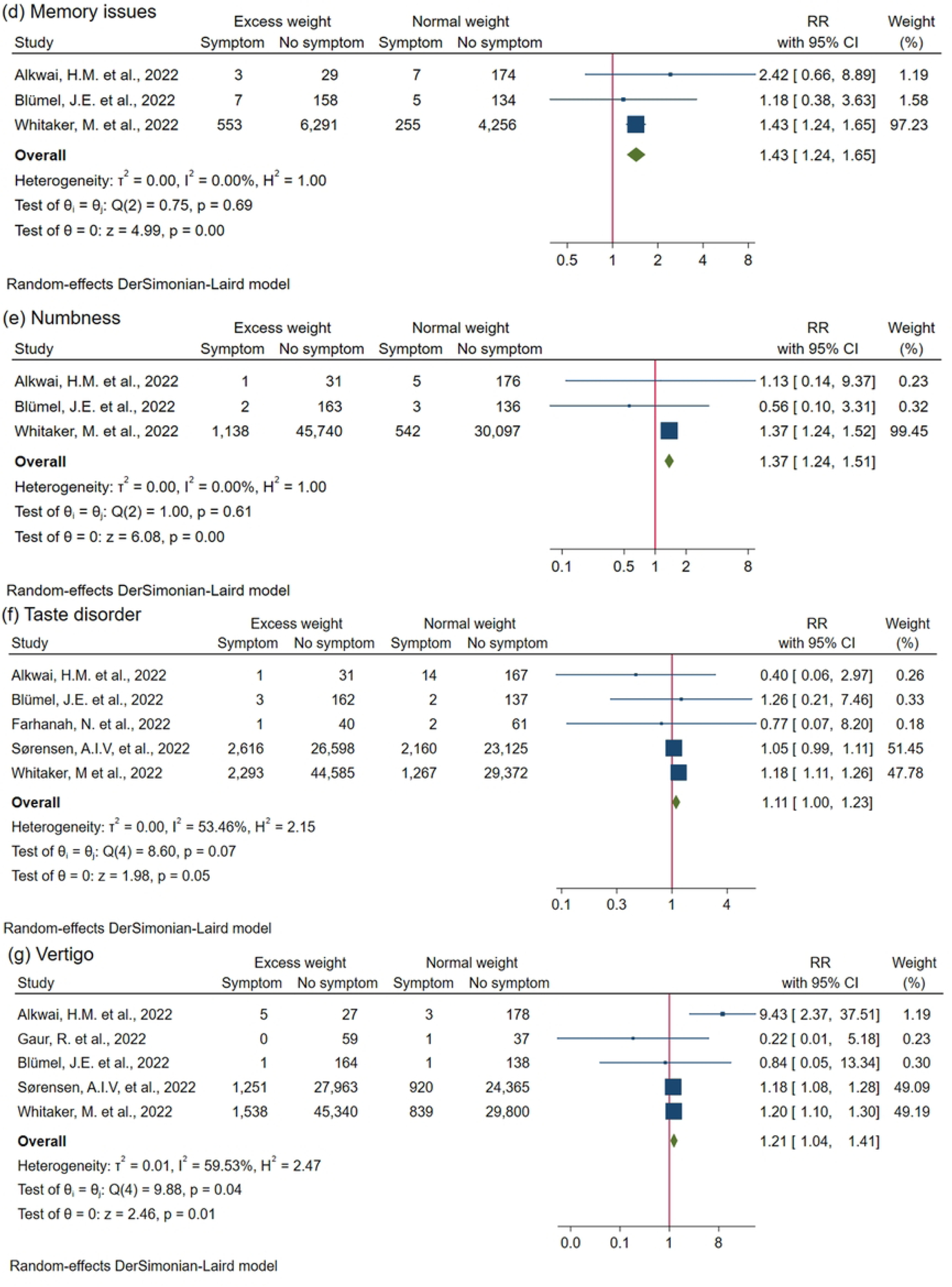
Forest plots of excess weight and risk ratio (RR) for neurological and neuropsychiatric symptoms (Continuation)

Four studies employed multivariate logistic regression and reported the risk of developing headache, smell disorder, memory impairment and taste disorder. These data were included in the meta-analysis. Pooled results revealed a significant association indicating that the risk of developing taste disorder was inversely associated with BMI (OR 0.93; 95%CI 0.88-0.98; *I*^2^=0,00). No significant associations were found for other evaluated symptoms (S2 Fig 2).

### Obesity and the Risk of Neuropsychiatric Symptoms

Our pooled risk analysis for obesity and neurological and neuropsychiatric symptoms (Figs 4a and 4b) showed that individuals with obesity are at significantly higher risk than those with normal weight or overweight (non-obesity control group) for a range of neurological symptoms: headache (RR = 1.41; 95% CI: 1.34–1.49; *I*^2^ = 3.39); numbness (RR = 1.61; 95% CI: 1.46–1.78; *I*^2^ = 0.00); smell disorder (RR = 1.15; 95% CI: 1.09–1.21; *I*^2^ = 7.55); taste disorder (RR = 1.21; 95% CI: 1.07–1.36; *I*^2^ = 63.57); and vertigo (RR = 1.44; 95% CI: 1.35–1.53; *I*^2^ = 0.00). No statistically significant association was found between obesity and the risk of developing PCC-related neuropsychiatric symptoms (S1 Fig 1b).

**Fig 4a.**
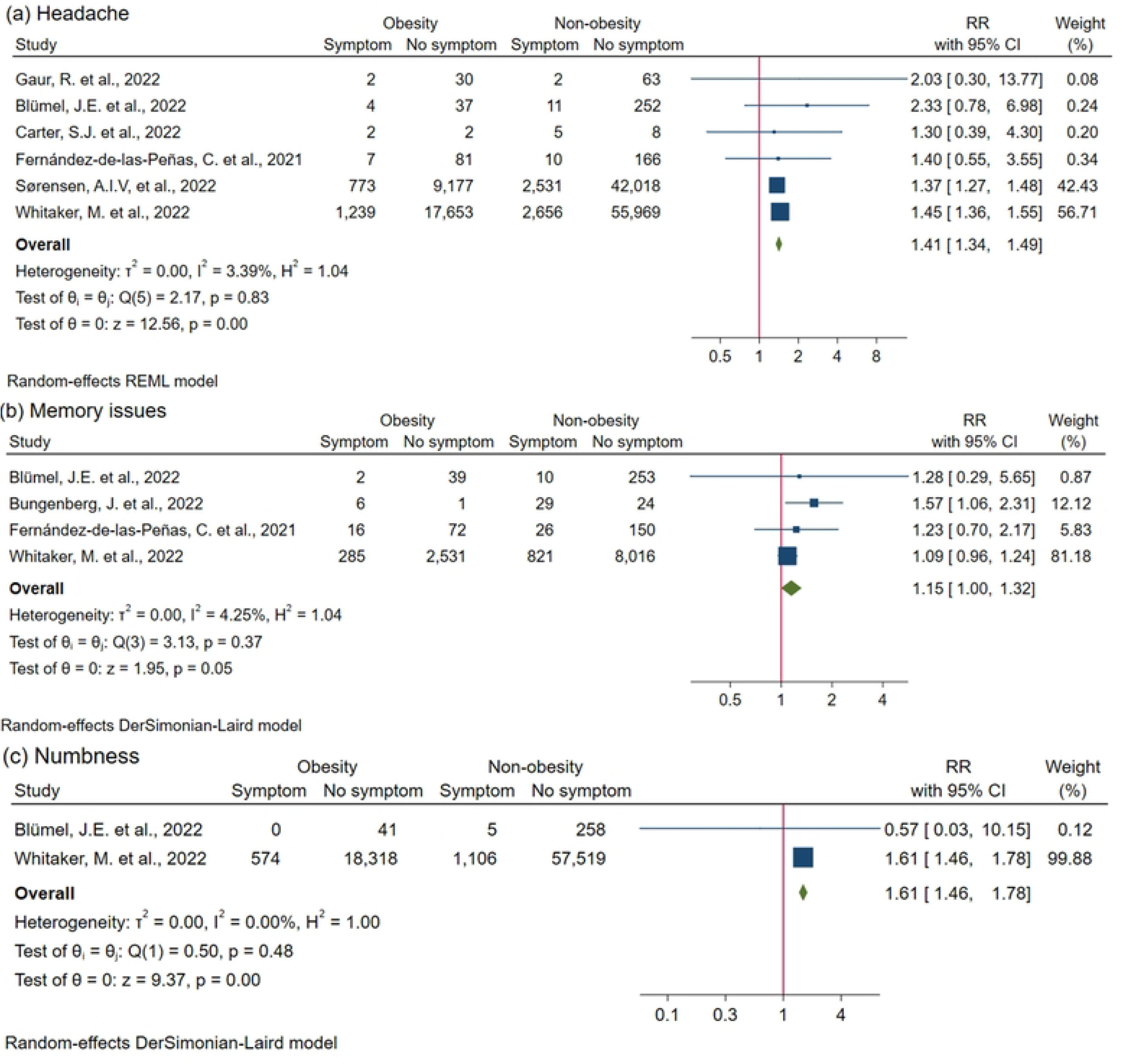
Forest plot of obesity and risk ratio (RR) for neurological and neuropsychiatric symptoms. ^a^ Applied a different BMI cut-off (obesity >25 kg/m^2^)

**Fig 4b.**
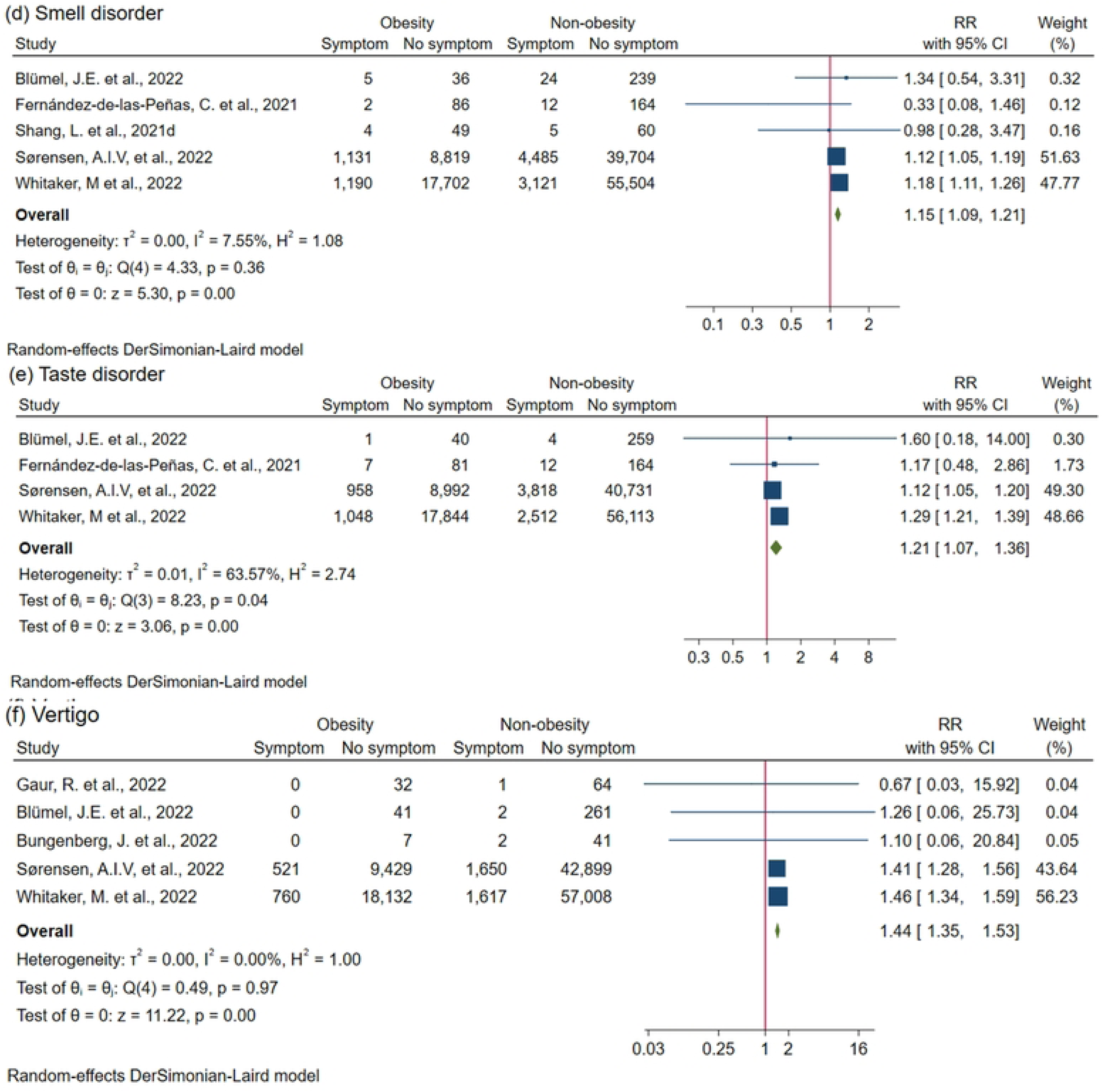
Forest plot of obesity and risk ratio (RR) for neurological and neuropsychiatric symptoms (continuation)

Substantial heterogeneity was observed in our meta-analysis for the non-significant association of obesity and anxiety (*I*^2^ = 70.5%) and sleep disorder (*I*^2^ = 61.94%) as well as for the statistically significant association between obesity and taste disorder (*I*^2^ = 69.5%). Meanwhile, no statistically significant heterogeneity was observed in other analyses where obesity was the exposure condition. Subgroup analyses could not be performed due to an insufficient number of studies evaluating the same outcome according to nutritional status.

### Risk-of-bias Assessment and Publication Bias

We assessed the risk of bias for each effect measure (frequency and OR). Results for studies reporting the frequency of PCC symptoms are presented in Fig 5; while findings for studies reporting OR are displayed in Supporting Information (S3 Fig 3). Among the 18 studies evaluated, seven were judged to have a high risk of bias due to self-reported exposure data[40, 43–45, 47, 52, 55]. The ROBINS-E tool indicated that no further evaluation of a study was required for studies with inappropriate measurement of exposure or outcome; therefore, this tool was applied only to the remaining 11 studies[32, 33, 41, 42, 46, 48–51, 53, 54].

**Fig 5:**
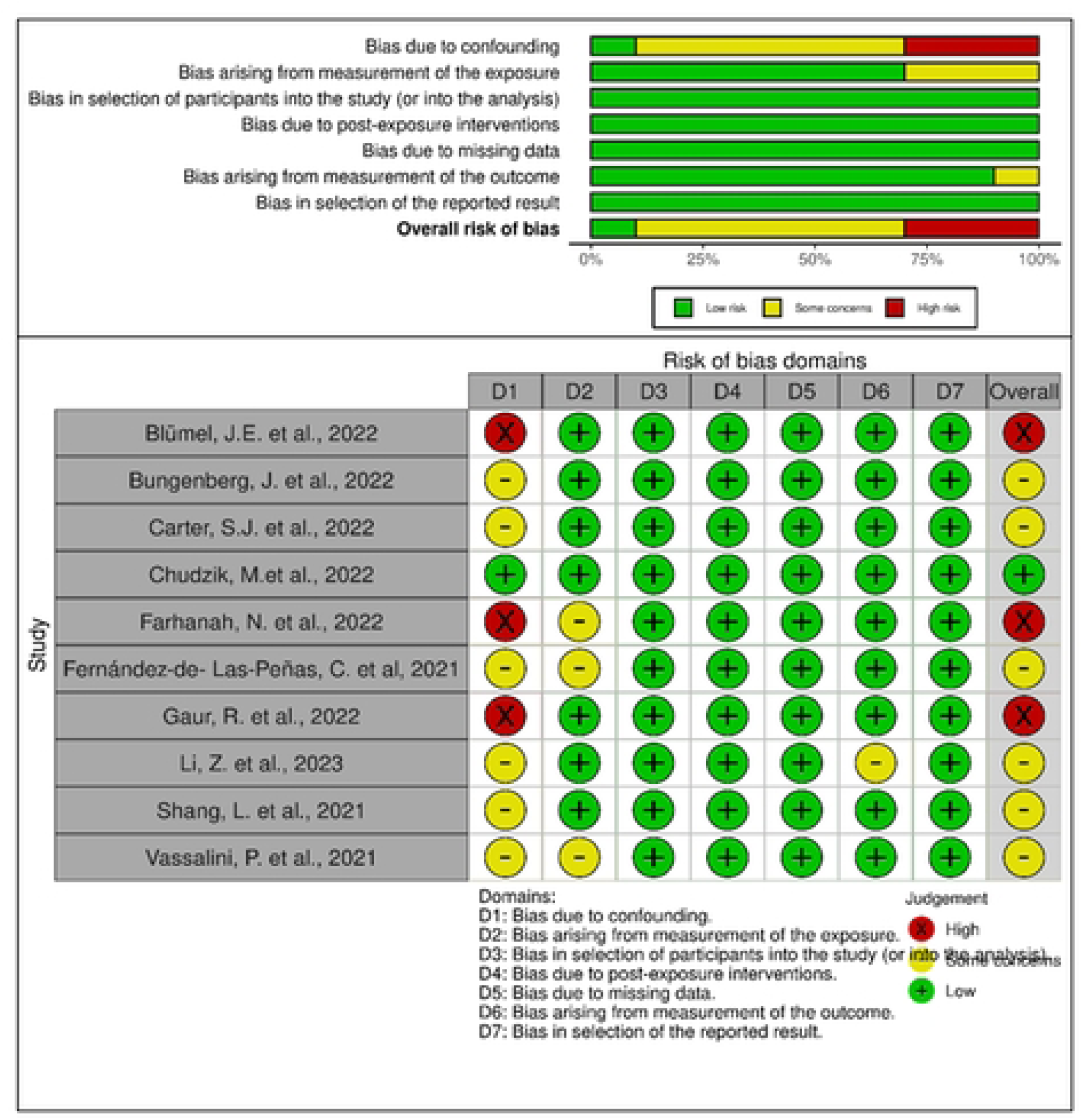
Summary plot and traffic light plot illustrating the risk of bias in the included studies, assessed using the ROBINS-E tool.

The domain most affecting the assessment was control for confounders (Domain 1 [D1]). Few studies employed appropriate designs (e.g., randomization or matching)[46, 51, 54] or analytical methods (e.g., stratification)[32, 48–50]; only one study had an overall low risk of bias[50]. Concerns were also noted regarding exposure measurement (Domain 2 [D2]), particularly when data were obtained from electronic medical records without reporting time-related information[51]. Visual assessment of the funnel plots did not imply asymmetry.

Findings from the GRADE assessment are presented in Table 3 and supporting information (S3 Table). The quality of evidence was downgraded from high to very low for three outcomes (headache, taste disorder, and vertigo) due to high risk of bias, imprecision of results, and publication bias. For the other outcomes, the quality of evidence was rated as low, having been downgraded from high due to risk of bias and publication bias.

**Table 3.**
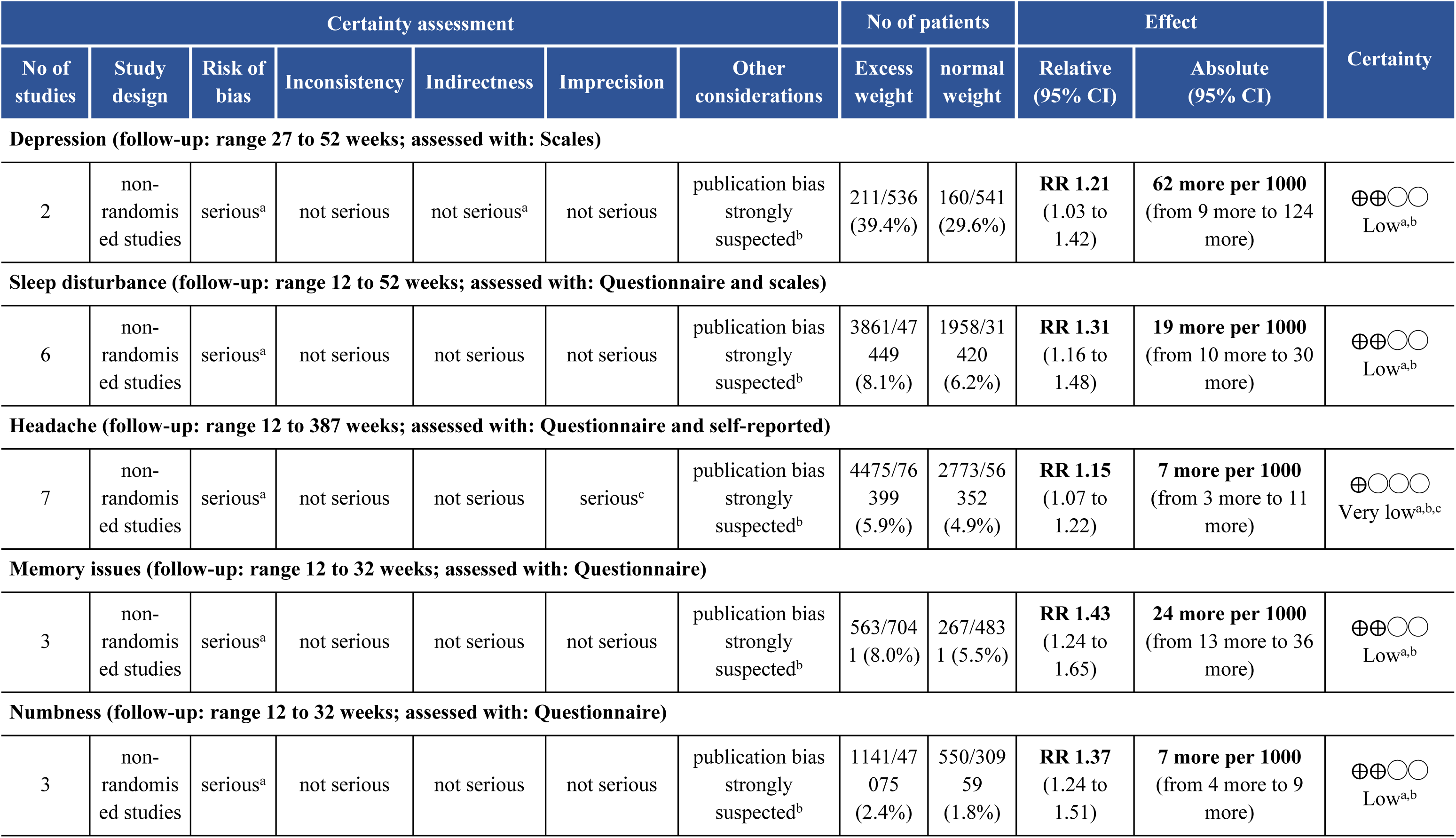
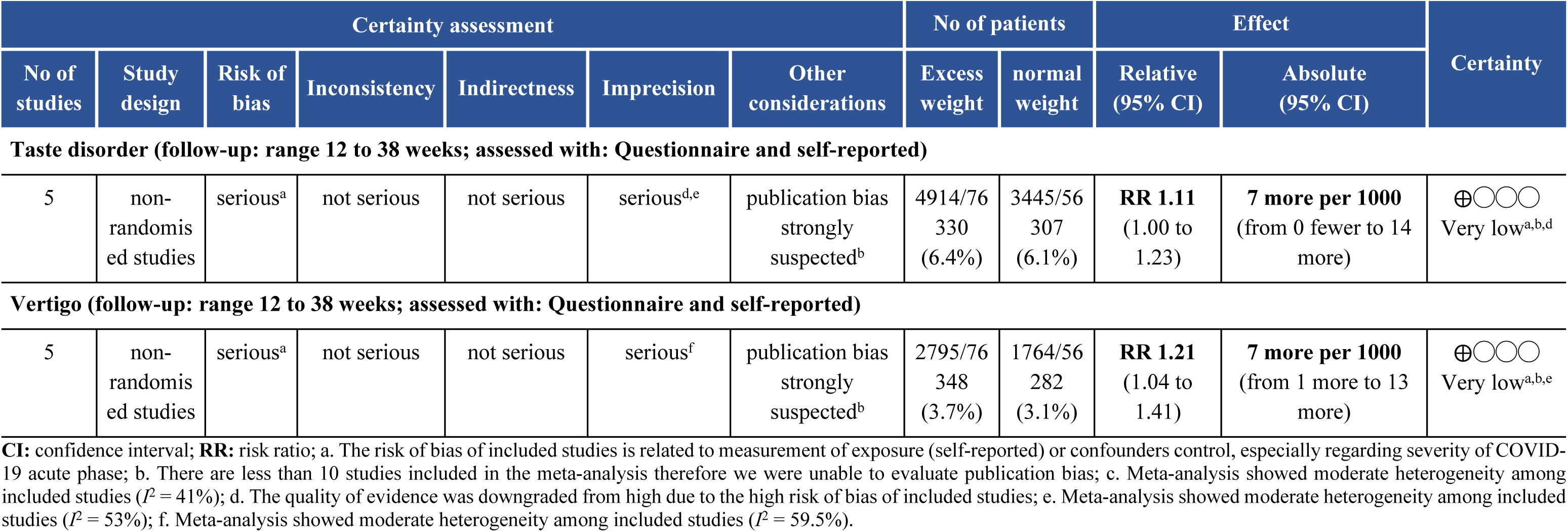
Assessment of the overall certainty of the evidence.

## Discussion

Although it has been suggested that individuals with increased BMI are at higher risk of PCC, the associations between EW and specific PCC related neurological and neuropsychiatric symptoms remained unclear. Our review reveals that EW is significantly associated with a range of persistent and PCC symptoms, including headache, vertigo, smell and taste disorder, sleep disturbance and depression. These findings suggest that EW markedly increases the risk of developing these symptoms that persist for more than 12 weeks after COVID-19 onset. Moreover, this study is opportune as we transition into the post-pandemic period facing the challenges of managing the co-occurrence of pandemics, including overweight/obesity, mental health issues, and the burden of PCC.

Increased BMI has been identified as a determinant of adverse outcomes during both the acute and chronic phases of COVID-19[16, 24, 26]. This contrasts with the role of sex in the disease course since the pattern of acute phase symptoms by sex is distinct from that of PCC, which tends to affect more females[18, 56]. Additionally, the chronic phase of COVID-19’s has been variously defined, with some studies indicating symptoms lasting for at least 4 weeks[10] and others extending to more than 12 weeks[5, 6]. Differentiating these time frames is crucial to distinguish between acute illness and potential sequelae of irreversible tissue damage, which may present with varying degrees of impairment[57]. In our review, we used the cut-off criterion to ensure that we focused on long-term or chronic outcomes of COVID-19. The mean follow-up time from illness onset in the included studies was 25.8 weeks (ranging from 12 to 52 weeks). A large systematic review[2] evaluated the prevalence of persistent symptoms among COVID-19 survivors at different follow-up periods. It identified sleep disorder and concentration difficulties as the most common symptoms at 3–6 months (24%, 95% CI: 8%–44%; 22%, 95% CI: 15%–31%, respectively), with sleep disorder being most prevalent at over 12 months. However, unlike our results, this review had a high degree of between-study heterogeneity and did not include BMI in subgroups analysis.

Long-term neurological symptoms significantly decrease the quality of life for individuals, a situation that might be exacerbated by a pre-exiting comorbidities[58]. Moreover, symptoms related to comorbidities often overlap with those of PCC, leading individuals with obesity to attribute their symptoms to their metabolic disease rather than to persistent manifestations of COVID-19[59, 60]. Additionally, overweight individuals also report health complaints, including headache and sleeplessness, associated to their subclinical disease state also known as suboptimal health status, which can be exacerbated by PCC symptoms[20]. The co-occurrence of these complex conditions can have deleterious effects, impairing people’s daily functioning and increasing the demand on healthcare systems. Our results highlight a significantly increased risk for persistent headache, vertigo, numbness and taste disorder in individuals with EW or obesity. This underscores the role of excessive body weight (fat deposits) in PCC. It is worth noting that patients with PCC have reported that head pain worsens with physical exercise while a decrease in physical activity coupled with increase in sedentary behaviour are generally known to contribute to weight gain[61],[62]. Moreover, taste disorders may influence unhealthy weight-related behaviours, such as consuming more palatable, ultra-processed foods high in salt, sugar, and additives. These foods can further impair gustatory function and exacerbate unhealthy symptoms[63, 64]. Vertigo which impairs daily functioning by increasing the risk of falls may also be associated with depression[65, 66]. Therefore, healthcare systems and public policies should focus on multidisciplinary rehabilitation services to address the long-term impacts of COVID-19 on survivors, specifically through concomitant personalized management of weight, neurological and neuropsychiatric issues.

The high prevalence of persistent complex concurrent symptoms may create a vicious circle between physical and neuropsychiatric symptoms, increasing the risk for depression[67–70]. A significant positive association between obesity and depressed mood has been observed across multiple COVID-19 periods, similar to findings in other coronavirus outbreaks[9, 14, 62]. During the COVID-19 pandemic, a systematic review of longitudinal studies identified obesity/overweight as a risk factor for depression (pooled RR = 1.2; 95% CI: 1.11-1.31). However, no significant association was found when obesity/overweight was self-reported (pooled RR = 1.03; 95% CI: 0.99–1.26)[71]. In this study, we observed a heightened risk of persistent depression among individuals with EW compared with those with normal weight. This findings aligns with Aminian *et al.*[72] who reported that the need for diagnostic test to assess neuropsychiatric problems (a *proxy* for symptoms) was significantly higher in individuals with increased BMI compared to those with normal weight. Although our pooled RR for EW and depression included data from two studies conducted with Asian populations[32, 40] featuring different BMI assessments (measured and self-reported) and BMI cut-offs (excess weight defined as BMI ≥ 23kg/m^2^ and obesity as BMI ≥25 kg/m^2^), both studies used a validated depression scale (PHQ-9), and the results were not heterogeneous.

It remains uncertain whether neurological and neuropsychiatric manifestations related to PCC are directly attributable to the virus itself or if they develop indirectly (such as, through an immune response or medical therapy). These symptoms may also involve both the central and peripheral nervous systems[14]. Additionally, the role of EW in the progression of COVID-19 is not fully understood, though it may be linked to an exaggerated inflammatory response or shared pre-existing genetic factors that these conditions share[73–75]. EW is also a recognized risk factor for various chronic conditions and the number of pre-existing comorbidities has been associated to the development of PCC symptoms up to two years after COVID-19 onset, in both hospitalized and non-hospitalized patients[76]. Our results underscore the importance of identifying at-risk individuals and highlight the need for timely personalized interventions for COVID-19 survivors. These findings could inform the development of predictive and preventive management plans for future waves of SARS-CoV-2 or other epidemics.

Our quality assessment results indicate that the included studies suffer from methodological issues, particularly in measuring exposure. Studies that classified individuals’ nutritional status based on self-reported data were found to have a high risk of bias, which contributed to downgrade in the overall quality of evidence of evaluated for the evaluated symptoms. During the height of the pandemic, online health surveys were often used when other research methods were infeasible[77]. However, significant issues can also arise from self-reported symptoms in studies that did not employ structured questionnaire or validated scales. Although screening tools have been proposed to identify people with PCC they often do not consider neuropsychiatric symptoms[78] or important cognitive symptoms[79] that significantly affect individuals’ quality of life. The observed moderate heterogeneity among studies evaluating headache, taste disorder and vertigo has led to imprecision in the results and the limited number of studies included in meta-analysis unable the assessment of publication bias, further contributing to downgrade of the overall quality of evidence for those outcomes from high to very low.

It is important to acknowledge certain limitations when interpreting the findings of our research. Most of the included studies were cross-sectional which limits the ability to determine cause and effect relationships. The inconsistency in naming PCC symptoms among studies resulted in a relatively low number of studies reporting the same symptom which also precluded subgroup comparisons between hospitalized and outpatient populations. We included studies that used non-validated questionnaires to assess symptoms, as it was necessary given that PCC is an emerging condition and validated screening tools have yet to be developed. Although the cut-off for study inclusion regarding time to follow-up (i.e., >12 weeks) allowed us to track long-term outcomes from COVID-19 and address a gap in the literature, the follow-up period ranged from 12 to 52 weeks post-infection, the varying risk of PCC symptoms throughout different follow-up periods was not investigated in subgroup analysis due to limited number of studies. Additionally, the inclusion of both obesity and non-obesity as comparisons groups led to the interchange of overweight individuals between exposure and control groups. Nevertheless, defining EW and obesity as exposure groups was crucial given the syndemic context. This approach enabled the evaluation of the risk of developing persistent symptoms among different levels of excessive fat and highlighted that both suboptimal health status (overweight) and illness (obesity) predisposed individuals to PCC-related neuro-symptoms. However, the inherent differences among these groups should be considered in health, disease prevention and diagnoses. Furthermore, the issues related to body fat assessment should be noted as BMI remains the most widely used screening tool for measuring adiposity. Although it is a simple and non-invasive measure often related to the gold standard fat assessment BMI does not accounts for the types or distribution of adipose tissue, nor does it for age and ethnic backgrounds[80]. Future research should focus on longitudinal evaluation of COVID-19 survivors incorporating objective assessment of symptoms and adiposity.

Despite these limitations, our review has several scientific and clinical strengths. Notably, the number of investigated outcomes allowed for a pioneering evaluation of the association of EW with both the physical and psychological long-term manifestations of COVID-19. Additionally, the comprehensive definition of PCC that we applied during our article search (to account for varied terminology), along with the inclusion of each neurological and neuropsychiatric symptom specified in our search strategy, ensured a thorough screening process. We also underscored the importance of addressing EW as a global health concern, given its interplay with neurological and neuropsychiatric manifestations, and infectious diseases such as COVID-19. The management of EW should be considered in the treatment of neurological and neuropsychiatric symptoms due to the significant impact that these combined conditions have on individuals’ health.

### Conclusions

Our systematic review and meta-analysis demonstrate that EW is significantly associated with an increased risk of developing post-COVID-19 neurological and neuropsychiatric symptoms, including headache, memory issues, numbness, smell and taste disorders, vertigo, depression, and sleep disturbance. These findings underscore the need for developing multidisciplinary rehabilitative strategies tailored to individual needs to improve care management and support the overall health of affected individuals. Our results provide evidence-based guidance for healthcare professionals and policymakers in managing PCC, and further support ongoing global efforts to understand its underlying mechanisms, epidemiology, and identification.

## Data Availability

All relevant data are within the manuscript and its Supporting Information files.

## Acknowledgements

We would like to thank the researchers who shared information and enabled more precise results in this study. We would also like to acknowledge students and collaborators of the PENSA research group at the University of Brasília and the Centre for Precision Health at Edith Cowan University for their support.

## Supporting Information

**S1 Table. Database search strategy.**

**S2 Table. Frequency of Post-COVID Condition neuropsychiatric symptoms of included studies that did not test for statistical differences according to nutritional status.** Excess weight group (BMI ≥ 25kg/m^2^); NW: Normal Weight group (BMI < 25kg/m^2^); Obesity group (BMI ≥ 30kg/m^2^); NOb: Non-obesity group (BMI < 30kg/m^2^).

**S1 Fig 1a: Forest-plots of excess weight and neuropsychiatric symptoms (non-significantly associations).**

**S1 Fig 1b. Forest-plots of obesity and neurological and neuropsychiatric symptoms (non-significantly associations)**

**S2 Fig 2. Forest-plot of the association of excess weight and the risk of neurological and neuropsychiatric symptoms.**

**S3 Fig 3. Traffic light plots of risk of bias of included studies that reported the risk of developing neuro-symptoms assessed by Robbins-e tool.**

**S3 Table 3. Assessment of the overall certainty of the evidence for studies thar compared the frequency of PCC neurological and neuropsychiatric symptoms among individuals with obesity and control groups**

